# The gut microbiome and rotavirus vaccine immunogenicity in rural Zimbabwean infants

**DOI:** 10.1101/2021.03.24.21254180

**Authors:** Ruairi C. Robertson, James A. Church, Thaddeus J. Edens, Kuda Mutasa, Hyun Min Geum, Iman Baharmand, Sandeep K. Gill, Robert Ntozini, Bernard Chasekwa, Lynnea Carr, Florence D. Majo, Beth D. Kirkpatrick, Benjamin Lee, Lawrence H. Moulton, Jean H. Humphrey, Andrew J. Prendergast, Amee R. Manges, SHINE Trial Team

## Abstract

**Background:** Oral rotavirus vaccine (RVV) immunogenicity is considerably lower in low-versus high-income populations; however, the mechanisms underlying this remain unclear. Previous evidence suggests that the gut microbiota may contribute to differences in oral vaccine efficacy.

**Methods:** We performed whole metagenome shotgun sequencing on stool samples and measured anti-rotavirus immunoglobulin A in plasma samples from a subset of infants enrolled in a cluster randomized 2×2 factorial trial of improved water, sanitation and hygiene and infant feeding in rural Zimbabwe (SHINE trial: NCT01824940). We examined taxonomic and functional microbiome composition using random forest models, differential abundance testing and regression analyses to explored associations with RVV immunogenicity.

**Results:** Among 158 infants with stool samples and anti-rotavirus IgA titres, 34 were RVV seroconverters. The median age at stool collection was 43 days. The infant microbiome was dominated by *Bifidobacterium longum*. The gut microbiome differed significantly between early (≤42 days) and later samples (>42 days) however, we observed no meaningful differences in alpha diversity, beta diversity, species composition or functional metagenomic composition by RVV seroconversion status. *Bacteroides thetaiotaomicron* was the only species associated with anti-rotavirus IgA titre. Random forest models poorly classified seroconversion status by both composition and functional microbiome variables.

**Conclusions:** RVV immunogenicity is low in this rural Zimbabwean setting, however it is not explained by the composition or function of the early-life gut microbiome. Further research is warranted to examine the mechanisms of poor oral RVV efficacy in low-income countries.

## INTRODUCTION

Rotavirus is the leading cause of diarrheal morbidity and mortality in children [1]. Although several oral rotavirus vaccines (RVV) are currently available globally, their efficacy varies substantially by population, limiting their impact [2]. Large efficacy studies show RVV efficacy of 85-98% against severe rotavirus gastroenteritis in high-income settings [3, 4], compared to only 48% and 39% in South Asia [5] and sub-Saharan Africa [6] respectively. The reasons for poor oral vaccine efficacy in low-income populations remain poorly understood, however intestinal factors may contribute [7-9].

The gut microbiota plays a critical role in the maturation of early-life immune function and intestinal development [10]. The composition of the early-life gut microbiota may also influence susceptibility to viral infections, since antibiotic treatment and segmented filamentous bacteria both reduce the pathogenicity of rotavirus in mice [11, 12]. Emerging evidence suggests that the early-life gut microbiota influences oral vaccine responses by modulating the immune and metabolic environments of the intestine [13]. In mice, antibiotic treatment impairs vaccine responses, an effect that is reversed following restoration of the microbiota [14]. In adults, narrow-spectrum antibiotics did not alter absolute anti-RV IgA titres following RVV, but did lead to a higher proportion of individuals displaying short-term IgA “boosting” (defined as a ≥2-fold rise in anti-RV titre 7 days post-vaccination) compared with placebo or broad-spectrum antibiotics [15]. Observational evidence from children in low-income settings has shown no consistent association between the gut microbiota and RVV efficacy in India [16], while RVV immunogenicity was associated with gut microbiota composition in cohorts in Pakistan [17] and Ghana [18]. In these settings, RVV seroconverters compared to non-seroconverters displayed a gut microbiota more similar to infants from a high-income setting.

We recently reported outcomes of a cluster-randomized trial of improved water, sanitation and hygiene (WASH) in rural Zimbabwe [19]. Among a subset of infants in whom anti-rotavirus IgA titres were measured, only 24% infants seroconverted following oral RVV administration. However, the WASH intervention significantly increased RVV seroconversion by 50% (19.6% in non-WASH arm versus 30.3% in the WASH arm) [20]. We hypothesised that the WASH intervention had reduced the burden of enteropathogens in these infants, thereby improving RVV immunogenicity. However, the WASH intervention had no effect on enteropathogen carriage [21] and enteropathogen burden was not associated with RVV immunogenicity [22]. In this follow-on study, we explore whether alterations in the composition of the gut microbiome are associated with RVV immunogenicity, by conducting whole metagenome shotgun sequencing of stool samples from infants around the time of RVV. Here we report on (i) the composition and function of the early-life gut microbiome in this rural LMIC setting and (ii) its association with RVV immunogenicity.

## MATERIALS AND METHODS

### Study population

We present a sub-study of the Sanitation Hygiene Infant Nutrition Efficacy (SHINE) trial [19], a 2×2 factorial cluster-randomized trial assessing the independent and combined effects of improved WASH and improved infant and young child feeding (IYCF) on stunting and anaemia (NCT01824940). Briefly, 5280 pregnant women in rural Zimbabwe were enrolled and cluster-randomized to standard-of-care, IYCF, WASH or combined IYCF+WASH, and children were followed up until 18 months of age. 1169 HIV-unexposed children were enrolled in a sub-study of the SHINE trial which involved longitudinal specimen collection (including plasma and stool) at 1, 3, 6, 12 and 18 months of age [23]. Sterile collection tubes were provided to mothers, who collected stool samples from their infants on the day of the study visit. Blood samples were collected from infants by trained nurses. Samples were transported by motorbike in cool boxes to field laboratories where they were aliquoted and stored at -80°C before subsequent transport to the central laboratory in Harare for ongoing storage. 158 infants with available plasma samples, oral rotavirus vaccine records and stool samples collected within 30 days of either vaccine dose were selected for analysis in this study. The SHINE trial was approved by the Medical Research Council of Zimbabwe and the Johns Hopkins Bloomberg School of Public Health Committee on Human Research. Written informed consent was obtained from all caregivers before enrolment in the trial.

### Sub-study population

For this study, infants were selected if they had at least one plasma sample collected post-rotavirus vaccination and an available stool sample collected at either 1 or 3-months of age, as per previous analyses [22]. We permitted a 30-day window of stool collection pre- or post-vaccination (either RVV dose), hypothesizing that the gut microbiome soon before, during or after vaccination may contribute to RVV immunogenicity. In order to assess the effect of this chosen 30-day time-window around vaccination, we conducted two sensitivity analyses: first, employing a restricted time-window to include those collected prior to 43 days of age (median age of sample collection); and second, restricting to samples collected within 14-days pre or post-RVV. Infants were excluded if they did not have vaccination data recorded or had not received at least one dose of RVV. If more than one sample was available per child, the stool sample collected closest to the first dose of RVV was chosen for analysis. 158 participants meeting these criteria were included in this study. 67 of these participants had previously been selected and had metagenome sequencing data generated as part of a separate investigation into longitudinal infant microbiota maturation from the same cohort [24]. The remaining 91 samples were selected at random for the current study, whilst oversampling seroconverters to increase statistical power, and processed in an identical manner.

### Rotavirus vaccination and anti–rotavirus Immunoglobulin A assay

Oral monovalent RVV (Rotarix™: GSK Biologicals) was introduced as part of the national vaccination programme in Zimbabwe in May 2014 and was administered to infants at 6 and 10 weeks of age. National vaccination coverage in 2015-2016 was 87-91% [25]. Vaccination was undertaken at local clinics as part of routine national vaccination programmes and was not overseen by the SHINE trial team. Vaccination dates were recorded by SHINE study staff by reviewing child health cards. Plasma anti–rotavirus immunoglobulin A (IgA) was measured on stored plasma samples by enzyme-linked immunosorbent assay (ELISA) using methods previously described [26]. The primary outcome was RVV seroconversion, defined as a post-vaccine plasma concentration of anti–rotavirus IgA ≥20 U/mL in infants who were seronegative (<20 U/mL) pre-vaccination [27]. Secondary outcomes included anti–rotavirus IgA titre and RVV seropositivity, defined as a post-vaccine titre ≥20 U/mL, regardless of pre-vaccine titre. We refer to these outcomes collectively as RVV immunogenicity.

### Stool metagenome sequencing

DNA was extracted from ∼200mg stool using the Qiagen PowerSoil Kit with bead beating. DNA quality was confirmed by spectrophotometry (SimpliNano) and quantified by fluorometry (Qubit). 1µg DNA was used as an input for sequencing library preparation following the Illumina TruSeq PCR-free library preparation protocol, using custom end-repair, adenylation and ligation enzyme premixes. Constructed libraries were assessed for quality of concentration (qPCR) and size (Tapestation 2200) prior to pooling. 48 barcoded samples were pooled in each sequencing run. Positive controls (ZymoBIOMICS) and DNA-free negative controls were included through all steps including DNA extraction and library preparation. One negative control was included in each sequencing pool. Whole metagenome sequencing was performed with 125-nucleotide paired-end reads using either the Illumina HiSeq 2500 or HiSeqX platforms at Canada’s Michael Smith Genome Sciences Centre, Vancouver, Canada.

*KneadData* was used with default settings to remove short reads (<60bp), duplicate reads, human and other non-microbial reads and to trim off adapters. Filtered sequencing reads were processed through the *MetaPhlan3* pipeline [28] with default settings to generate compositional data. *HUMAnN3* was used with default settings using the UniRef90 database to generate functional annotations (enzyme commission (EC) annotations and pathways) [29]. Compositional and functional data were filtered using a minimum threshold of >0.1% and >0.0001% relative abundance, respectively. Taxa were included in the analyses if present at a minimum threshold of 5% prevalence within the entire dataset.

A median of 8.1 ± 3.0 million quality filtered reads were produced per sample. Sixteen negative controls were also sequenced with mean 734 quality-filtered reads (range = 149 to 11,432, SD = 3462). Following filtering of annotatable reads, 139 species were included in the final analysis, of which 4 were classified as co-abundance gene groups metagenomic assemblies (CAGs) for which no culture-derived representative exists.

### Statistical Analyses

All data were analysed using R (v.4.0.2). Beta-diversity was estimated using the Bray-Curtis dissimilarity index. Alpha diversity was assessed using the Shannon index and the number of observed species. Differential abundance analysis was performed by Wilcoxon rank sum test and Analysis of Composition of Microbiomes (*ANCOM*) *[30]*. Random Forest machine learning models were performed using the *randomForest* package (*SIAMCAT* package [31]) in which five-fold cross-validation was performed with 100 iterations. Microbiome regression analyses were performed using the *MaAsLin2* package. Seven covariates were chosen for adjustment in regression models and included age at stool sample collection, birthweight, exclusive breastfeeding status (recorded at 3 months old), sex, WASH randomized trial arm, delivery mode and length-for-age Z-score (LAZ) around time of vaccination. These covariates were chosen based on biological plausibility and previous evidence of their independent influence on RVV immunogenicity in the same cohort [32]. Multiple comparisons were tested using Benjamini-Hochberg false discovery rate (FDR; q-value <0.05).

## RESULTS

### Study cohort

Among 5280 women enrolled in the SHINE trial, there were 3989 live-born, HIV-unexposed infants. Of these, 882 children had RVV immunogenicity measured and 158 of these had stool samples collected within 30 days of either vaccine dose and with valid metagenome sequencing data. **Table 1** outlines the primary characteristics of the 158 participants in this analysis. Of these 158 participants, 115 had seroconversion data available as a primary outcome (n=81 non-seroconverters, n=34 seroconverters; **Figure 1a**), whilst secondary outcomes (seropositivity status and IgA titres) were available for all 158 infants (n=113 seronegative, n=45 seropositive).

**Table 1.**

|  | Seroconversion |  |  | Seropositivity |  |  |
| --- | --- | --- | --- | --- | --- | --- |
|  | Non-seroconverters | Seroconverters | P value | Seronegative | Seropositive | P value |
|  | N=81 | N=34 |  | N=113 | N=45 |  |
| Female, % | 61.7 | 55.9 |  | 59.3 | 48.9 |  |
| Birthweight, kg (sd) | 3.17 (0.48) | 3.06 (0.55) | 0.303 | 3.11 (0.49) | 3.06 (0.50) | 0.564 |
| Low birthweight, % | 4.94 | 11.8 |  | 7.96 | 8.89 |  |
| Vaginal delivery, % | 96.3 | 97.1 |  | 95.6 | 95.6 |  |
| Institutional delivery, % | 91.2 | 94.1 |  | 93.8 | 95.6 |  |
| Exclusively breastfed (at month 3), % | 90.0 | 97.1 |  | 90.1 | 95.5 |  |
| Concurrent OPV vaccination, % | 95.7 | 93.8 |  | 96.5 | 92.1 |  |
| Age at stool sample, days [IQR] | 38 [34; 66] | 37 [33; 41] | 0.222 | 45 [35; 78] | 38 [34; 59] | 0.034 |
| Age at first RV dose, days [IQR] | 46 [43; 54] | 44 [42;45] | 0.002 | 45 [42; 50] | 44 [42; 45] | 0.019 |
| Age at second RV dose, days [IQR] | 82 [74; 100] | 77 [75;82] | 0.037 | 78 [72; 93] | 77 [75; 83] | 0.669 |
| LAZ at 1 month visit (sd) | -0.88 (1.09) | -0.80 (1.43) | 0.783 | -1.00 (1.19) | -0.73 (1.33) | 0.267 |
| LAZ at 3 month visit (sd) | -0.90 (1.07) | -0.84 (1.19) | 0.786 | -0.94 (1.13) | -0.88 (1.25) | 0.791 |
| WASH trial arm, % | 42.0 | 47.1 |  | 41.6 | 51.1 |  |
| Born in RV season, % | 33.3 | 44.1 |  | 29.2 | 37.8 |  |
| Household size | 5 [4; 6] | 5 [4; 6] | 0.126 | 5 [4; 6] | 5 [4; 6] | 0.298 |
| Mother age at baseline, years (sd) | 27.9 (5.8) | 27.3 (6.1) | 0.659 | 27.6 (6.0) | 27.2 (6.5) | 0.787 |
| Mother height at baseline, cm (sd) | 160.7 (6.2) | 160.7 (6.0) | 0.983 | 160 (5.9) | 161 (6.2) | 0.722 |

**Fig 1.**
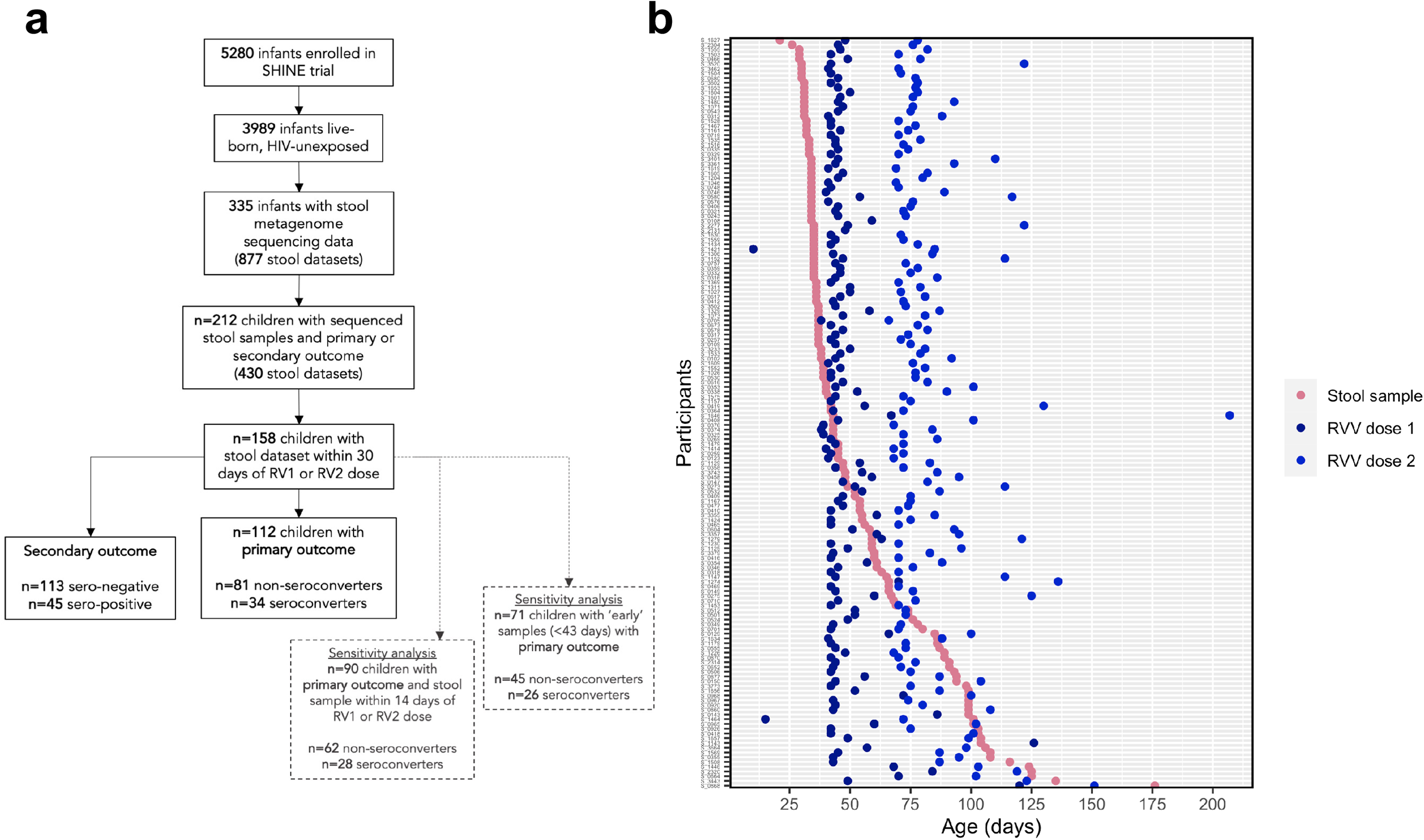
Study flow. (a) Flow diagram of stool samples included in the analysis originating from the SHINE trial. (b) Timing of stool samples included in the analysis and associated RVV doses.

The median age at first dose of RVV was 44 days [interquartile range (IQR): 42-49]. The median age at stool sample collection was 43 days (IQR: 35-68; **Figure 1b**). To test for and account for age-associated changes in microbiota composition, analyses were also split by early (≤ 42 days) and late (> 42 days) stool samples.

### Microbiome alpha and beta diversity associations with RVV immunogenicity

PERMANOVA analysis identified a small but significant difference in beta diversity as measured by Bray-Curtis distances by seroconversion status (P = 0.035; R^2^ = 0.014; **Fig. 2a**), however this was largely explained by data dispersion (homogeneity of dispersion test; P = 0.035). When split by median age of sample collection (43 days), there was a significant difference in Bray-Curtis distances between early and late samples (P <0.001; R^2^ = 0.025; **Fig. 2b**). Beta diversity analyses did not indicate a significant difference between samples collected immediately prior to or after the first dose of RVV in samples collected in the first 60 days of life (**Fig. S1a**), suggesting that there was no significant difference of the vaccine itself on gut microbiome beta diversity. No significant difference was observed between infants randomized to WASH or non-WASH trial arms (**Fig. S1b)**.

**Fig. 2.**
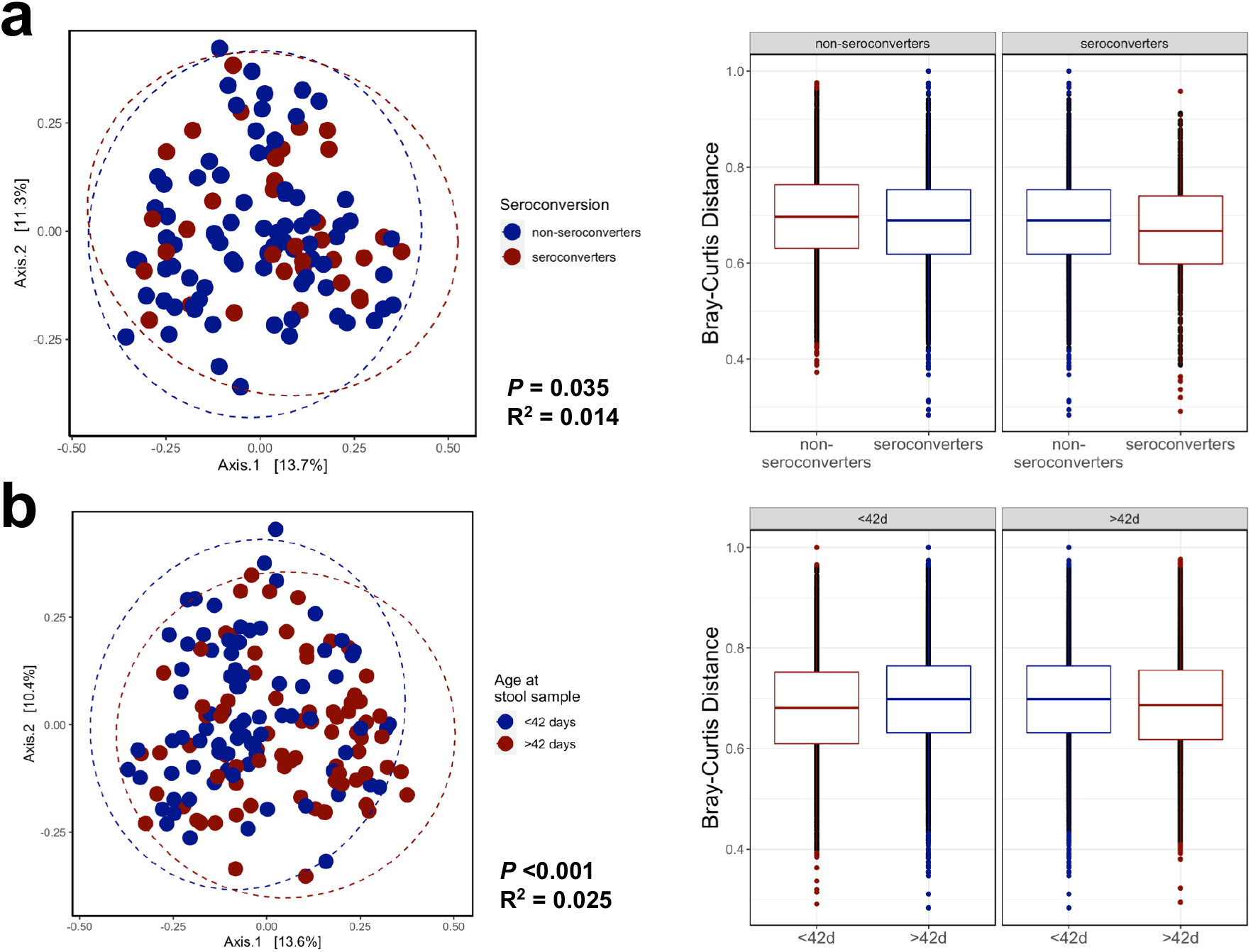
Beta diversity analysis by seroconversion status and early vs late samples. Principal coordinates analysis (PCoA) and associated Bray-Curtis distances to assess beta diversity of seroconversion status (a) and early vs late samples (b) as assessed using PERMANOVA analysis.

In sensitivity analyses, when restricting to samples collected within 14 days of either vaccine dose (n=62 non-seroconverters, n=28 seroconverters), there was a significant difference in Bray-Curtis distances by seroconversion status (P = 0.011; R^2^= 0.022; **Fig. S1 c**), which was partly explained by data dispersion (P = 0.03). However, there was no significant difference in Bray-Curtis distances by seroconversion status when restricted to early samples (≤ 42 days; n=79 samples; P = 0.061; R^2^ = 0.024). In analysis of secondary outcomes, beta diversity differed significantly by seropositivity status (P = 0.008; R^2^ = 0.012; **Fig. S1 d**) in the total dataset, which was not explained by dispersion (P = 0.067), but not when restricted to samples collected before ≤ 42 days (P = 0.113; R^2^ = 0.019).

Alpha diversity, as measured by the Shannon index and number of observed species, did not significantly differ by seroconversion (Wilcoxon test: P = 0.369 & P = 0.260 respectively; **Fig. 3a**) and was not associated with anti-rotavirus IgA titre in simple linear regression (*P* = 0.306; **Fig. 3b**). As expected, there were significant differences in alpha diversity between early and late samples (P = 0.032).

**Fig. 3.**
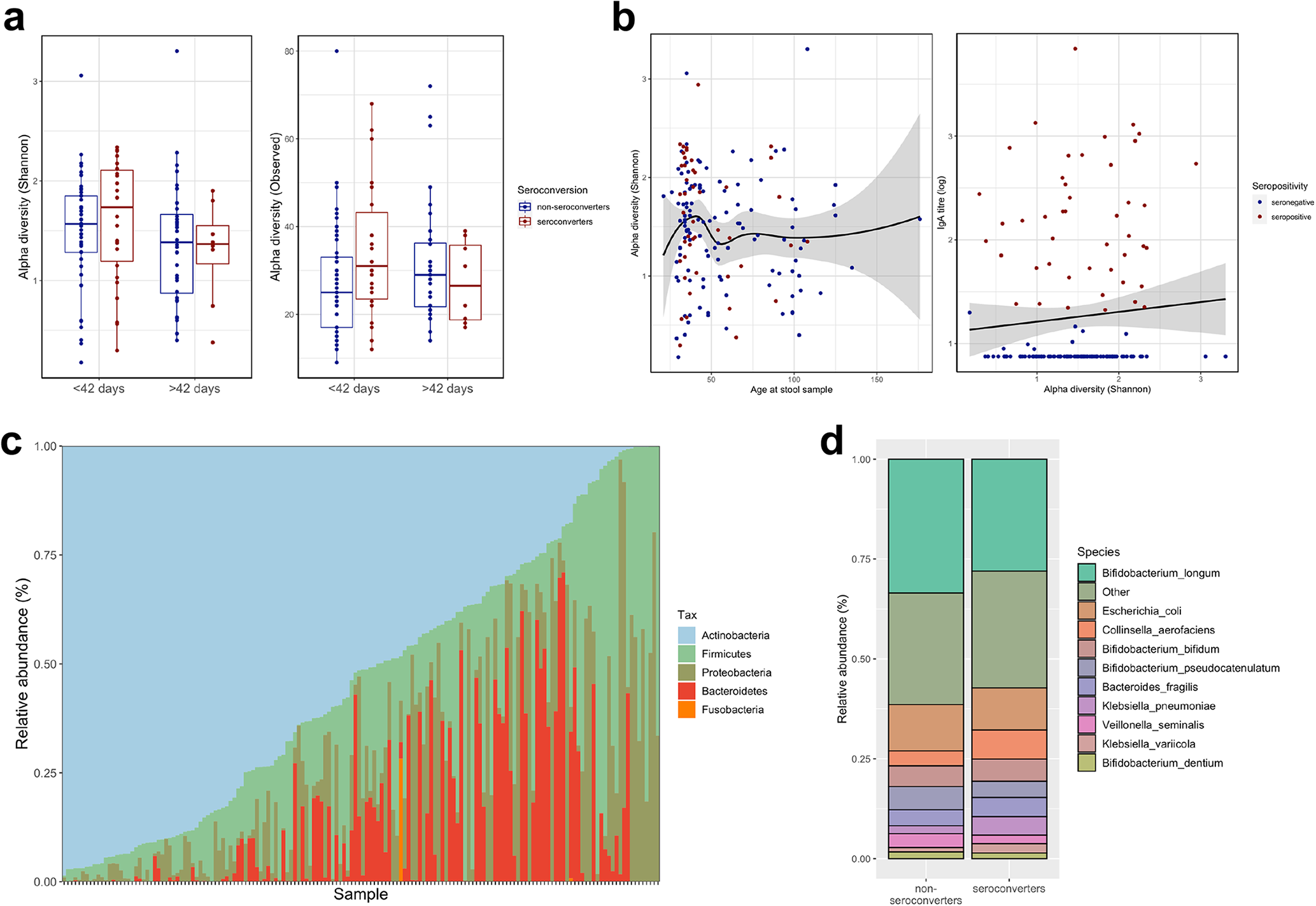
Alpha diversity and taxonomic composition. (a) Alpha diversity, as assessed using the Shannon index and number of observed species, between serconverters and non-seroconverters. (b) Associations between alpha diversity, age at stool sample collection and RV IgA titre. (c) Phylum composition of entire dataset. (d) Species composition in seroconverters vs non-seroconverters.

In sensitivity analyses, alpha diversity also did not differ by seroconversion status or seropositivity status when restricted to early samples (P = 0.333 and P = 0.260, respectively) or when restricted to stool samples collected within 14 days of either RVV dose (data not shown). There was also no significant difference in alpha diversity by seropositivity status (P = 0.18; **Fig. S2a-b**) or by randomized WASH arms (**Fig. S2c)**.

### Infant microbiome taxonomic composition and association with RVV immunogenicity

At the phylum level, samples were dominated by Actinobacteria (**Fig. 3c**). *Bifidobacterium longum* was the most abundant species in the entire dataset comprising a median 23% relative abundance, ranging from 0% to 97% across all samples. *Escherichia coli, Collinsella aerofaciens, B. bifidum* and *B. pseudocatenulanum* were the next most highly abundant across all datasets. Differential abundance analysis showed no significant differences in relative abundances of any taxa by seroconversion status (Wilcoxon ranked sum test, q > 0.05; **Fig. 3d**). This negative result was confirmed by the more stringent ANCOM model, which included covariate adjustment for age at stool sample collection, birthweight, exclusive breastfeeding status, sex, trial arm, delivery mode and LAZ around vaccination. Six species were significantly differentially abundant between early and late samples (Wilcoxon, q < 0.05: *B. longum, Streptococcus mitis, Bacteroides ovatus, Colinsella aerofaciens, Staphylococcus hominis, Streptococcus pneumoniae*; **Fig. 4a** and **S2e**) which was confirmed by ANCOM following covariate adjustment. In analysis of secondary outcomes, no taxa were significantly different by seropositivity status (**Fig. S2f**). In sensitivity analyses, when restricted to early samples or restricted to samples collected within 14 days of either vaccine dose, no taxa were significantly differentially abundant when compared by seroconversion or seropositivity statuses. *Collinsella aerofaciens* was the only species significantly different in relative abundance between WASH and non-WASH infants in this sub-cohort (**Fig. S2f**).

**Fig. 4.**
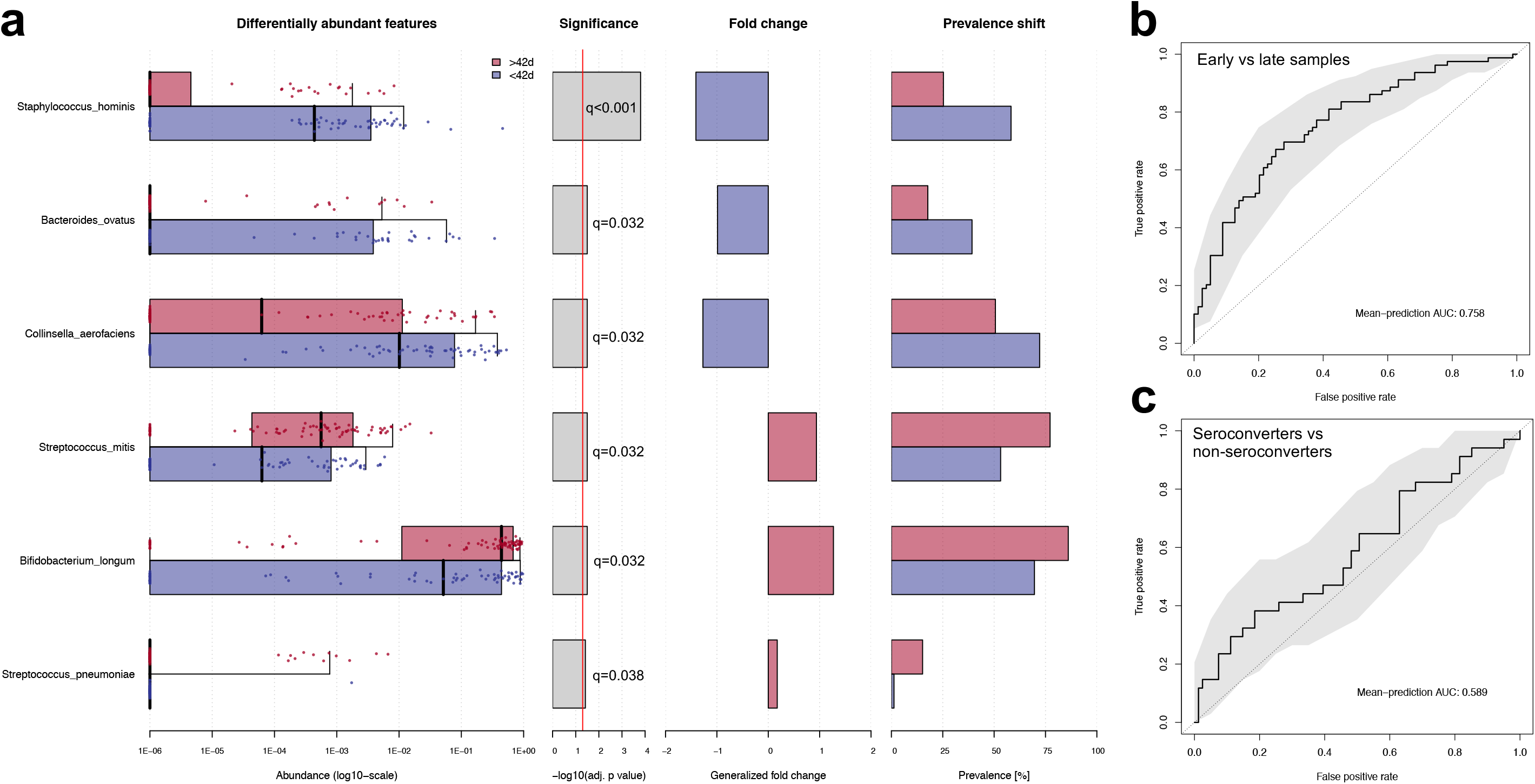
Differential abundance analysis and machine learning models of taxonomic composition. (a) Species that differ significantly in relative abundance between early (≤ 42 days) and late (> 42 days) samples as assessed by Wilcoxon ranked sum test (q value < 0.05). Random Forest machine learning classification model of (a) early vs late samples and (b) seroconverters vs non-seroconverters using all microbiome species.

Random Forest models were also applied to the datasets to test whether microbiome features could predict seroconversion and age. Early and late samples were strongly classified by the Random Forest model (receiver operating curve area under the curve (AUC) 0.758; **Fig. 4b**). Random Forest regression of age produced moderately strong models (*P* = 0.001; R^2^ = 0.152; 14% explained variation). However, random Forest poorly classified seroconversion status (AUC 0.589; **Fig. 4c**). In analysis of other secondary outcomes, random forest also poorly classified seropositivity (AUC 0.564; **Fig. S3a**) and WASH trial arm (AUC 0.5; **Fig. S3b**).

Regression models were built (MaAsLin2) to identify associations between gut microbial taxa and age or vaccine titres. Following FDR correction and adjustment for covariates, *Bacteroides thetaiotaomicron* was the only species significantly associated with IgA titre (P = 0.008). This effect held in sensitivity analyses using samples collected within 14 days of either vaccine dose (P = 0.053) whilst also indicating that *Slackia isoflavoniconvertens* was also associated with IgA titre in the restricted sensitivity dataset (P = 0.049).

### Infant microbiome functional composition and association with RVV immunogenicity

Metagenomic data were also analysed to assess microbiome functional pathways associated with age and RVV responses. Wilcoxon tests identified no functional pathway or EC that differed significantly by seroconversion status. Random Forest models of microbiome functional pathways also poorly classified seroconversion status. Two metagenomic pathways (*PWY*−*5994: palmitate biosynthesis I (animals and fungi* and *PWY*−*1042: glycolysis IV (plant cytosol)*) and 153 ECs were differentially abundant between early and late samples (**Fig. 5**). Random Forest models performed moderately well in classifying early and late samples based on metagenomic pathways (AUC 0.641; **Fig. S3c**). No EC or metagenomic pathway was significantly associated with IgA titre. In sensitivity analyses of early samples or those collected within 14 days of either vaccine dose, no EC or metagenomic pathway was associated with seroconversion and random forest classification was poor. No significant functional differences were observed by seropositivity status.

**Fig. 5.**
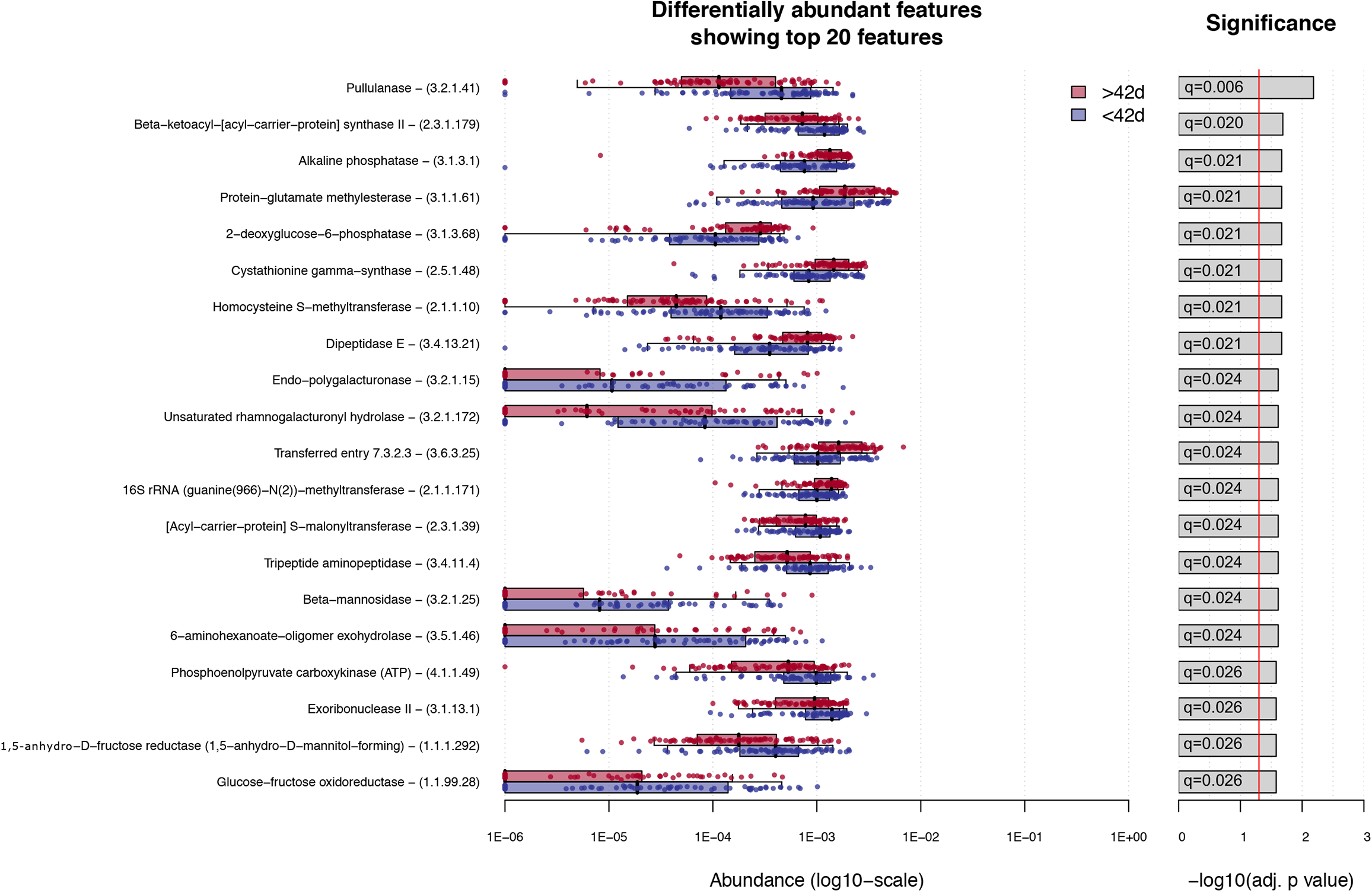
Functional metagenomic analysis. Enzyme commission annotations that differ significantly in relative abundance between early vs late samples (Wilcoxon rank sum test; q value < 0.05; top 20 ECs shows as ranked by q-value).

## DISCUSSION

Here, we report on the stool metagenomes of 158 infants <6 months of age from a rural Zimbabwean birth cohort and their association with RVV immunogenicity. Similar to other low-income settings, RVV was poorly immunogenic in this cohort (24% seroconversion). The reasons for this are unclear, but dynamic changes in the microbiota at this age may influence mucosal immune responses to an oral vaccine. However, we demonstrate here that there is no distinct gut microbiome signature that distinguishes RVV seroconverters from non-seroconverters. Hence, poor vaccine seroconversion in this population does not appear to be explained by the gut microbiome.

A handful of previous studies have demonstrated conflicting evidence for the association between the infant gut microbiome and oral RVV efficacy. A previous analysis of 170 infants in south India, using 16S amplicon sequencing, failed to find an association between the commensal infant gut microbiome and RVV seroconversion [16]. In the same study, however, RVV seroconverters were more likely to harbour at least one bacterial enteropathogen than non-seroconverters. Conversely, two separate studies in Ghana (n=20) [18] and Pakistan (n=78) [17], employing HITChip microarrays rather than next-generation sequencing, both demonstrated that the gut microbiome of RVV seroconverters differed significantly to non-seroconverters. Both of these studies also reported that the gut microbiome of seroconverters versus non-seroconverters were more similar to infants from a high-income setting in the Netherlands. However, the taxa associated with RVV seroconversion differed between countries. Higher relative abundances of Clostridium cluster XI and Proteobacteria were associated with seroconversion in Pakistan, whilst higher *Streptococcus bovis* and lower Bacteroidetes were associated with seroconversion in Ghana.

Due to the methodological differences in sequencing and the geographic differences in the cohorts, it is difficult to draw comparisons with these previous studies. Geographic setting has a major influence on gut microbiome composition and inter-individual variation between children across geographical settings is greater than in adults [33]. The results presented here, using gold-standard metagenome sequencing, failed to find a clear infant microbiome signature that distinguished RVV seroconverters from non-seroconverters. Previous evidence shows that the strongest contributor to infant gut microbiome composition in early-life is exclusivity of breastfeeding [34], which was extremely high in this cohort (>80%) due to study interventions designed to promote early and exclusive breastfeeding. Hence, any gut microbiome signatures associated with RVV seroconversion may be outweighed by the influence of breastfeeding and other exposures on the early-life microbiome.

We recently reported that enteropathogen carriage was not associated with RVV seroconversion in this cohort [22]. The results presented here extend these observations by showing that neither commensal nor pathogenic enteric microbes influenced RVV seroconversion. Oral RVV is taken up in the small intestine, whereas the gut microbiome and enteropathogen carriage measured in stool largely reflect the colonic intestinal environment. This may partly explain why we observed no association between the stool microbiota and RVV seroconversion in this cohort. Future studies examining the small intestinal microbiota may provide greater insight into the potential interaction between the commensal gut microbiome and oral vaccines. It remains unclear what biological factors contribute to poor RVV immunogenicity in low-income settings. Improved WASH enhanced RVV immunogenicity by 50% in this cohort [20], yet had no impact on enteropathogen carriage [21] or diarrhea [19]. In this subset of infants, WASH similarly had no significant impact on the early-life gut microbiome, suggesting that the effect of WASH on RVV immunogenicity is not driven through the gut microbiome. Hence, the causal pathway linking improved WASH with improved RVV seroconversion remains unexplained and complex.

This study benefited from a well-characterized cohort of infants from a large, community-based, cluster randomized trial. RVV seroconversion, measured in a subset of infants, was 23.7% rate despite high vaccine coverage, demonstrating its suitability as a representative infant population with low RVV immunogenicity. Within this subgroup, we have previously investigated and reported on a number of environmental factors associated with RVV seroconversion including enteropathogen carriage, WASH, household factors, birthweight and nutritional status [20, 22, 32]. Hence the data presented here, albeit negative, add to the evidence base of RVV immunogenicity in a unique, well-characterised cohort by contributing unique insight into the intestinal milieu and its association with oral vaccine efficacy. We employed whole metagenome shotgun sequencing, which has not previously been extensively conducted in low-income settings, allowing us to examine both taxonomic and functional microbiome variables in relation to RVV seroconversion. It is also one of the very few high-resolution, metagenomic datasets from infants in a rural, non-industrialized setting.

The study is limited by its relatively small sample size, which is partly due to the low seroconversion rate in this population, the number of stool samples available for sequencing analysis and the criteria of sample selection within a 30-day time-window close to vaccination. This arbitrary time-window was based on the hypothesis that the intestinal environment around the time of vaccination may alter vaccine efficacy and was chosen to complement our previous analyses in this same cohort using the same cut-off around vaccination [22]. The study is limited by this chosen time-window which may lead to confounding effects of age on any potential gut microbiome associations. To mitigate this, we adjusted for age in all regression models and also performed sensitivity analyses using a restricted 14-day time window and analysing a subset of ‘early’ samples collected before 43 days of age (median age of sample set). Secondly, the outcomes assessed in this study (seroconversion and seropositivity) may not be accurate correlates of vaccine protection. We focussed on seroconversion as a primary outcome as this depends on both pre and post-vaccine titres and also included seropositivity as a secondary analysis. However, as we and others have reported [32], natural rotavirus infection prior to vaccination may confound IgA titres which limits both seroconversion and seropositivity in particular as an outcome. Furthermore, seroconversion and seropositivity are not perfect correlates of protection against rotavirus protection [35]. Thirdly, due to the difficulties in obtaining viral reads from stool metagenomic data, we were unable to determine the impact of enteroviruses on RVV seroconversion, which has been hypothesized previously to be associated with RVV immunogenicity [16]. Specific analysis of the infant virome may shed more light on RVV underperformance.

In conclusion, we found no clear stool microbiome signature associated with oral RVV immunogenicity in rural Zimbabwean infants. Further research into other intestinal and additional biological pathways that explain poor oral vaccine immunogenicity in low-resource settings is warranted.

## Data Availability

Individual level data will be uploaded by the trial team as individual participant data with an accompanying data dictionary at http://ClinEpiDB.org in mid 2021. This platform is charged with ensuring that epidemiological studies are fully anonymised by removing all personal identifiers and obfuscating all dates per participant through application of a random number algorithm to comply with the ethical conduct of human subjects research. Researchers must agree to the policies and comply with the mechanism of ClinEpiDB to access data housed on this platform. Prior to that time, the data are housed on the ClinEpiDB platform at the Zvitambo Institute for Maternal and Child Health Research and available upon request from Ms. Virginia Sauramba.

## FUNDING

This work was supported by the Wellcome Trust [203905/Z/16/Z to JAC, 206455/Z/17/Z to RCR and 093768/Z/10/Z and 108065/Z/15/Z to AJP]. The SHINE trial was funded by the Bill and Melinda Gates Foundation [OPP1021542 and OPP113707]; UK Department for International Development (UK Aid); Swiss Agency for Development and Cooperation and US National Institutes of Health [2R01HD060338-06]. The study funders approved the trial design, but were not involved in data collection, analysis, interpretation, or manuscript preparation.

## CONFLICT OF INTEREST

The authors declare no conflicts of interest

## ACKNOWLEDGEMENTS

The authors thank all the mothers, babies and their families who participated in THE SHINE trial. The authors gratefully acknowledge the leadership and staff of the Ministry of Health and Child Care in Chirumanzu and Shurugwi districts and Midlands Province (especially environmental health, nursing and nutrition) for their roles in operationalization of the study procedures. The authors acknowledge the Ministry of Local Government officials in each district who supported and facilitated field operations. The authors also acknowledge Monica M. McNeal for her invaluable support in setting up the anti-rotavirus IgA assay.

**All authors attest they meet the ICMJE criteria for authorship**

## Notes

### Competing Interest Statement

The authors have declared no competing interest.

### Clinical Trial

NCT01824940

### Author Declarations

The Medical Research Council of Zimbabwe and the Institutional Review Board of the Johns Hopkins Bloomberg School of Public Health approved the study protocol.

