## Supplementary figures (S1-S3) for "The gut microbiome and rotavirus vaccine immunogenicity in rural Zimbabwean infants"

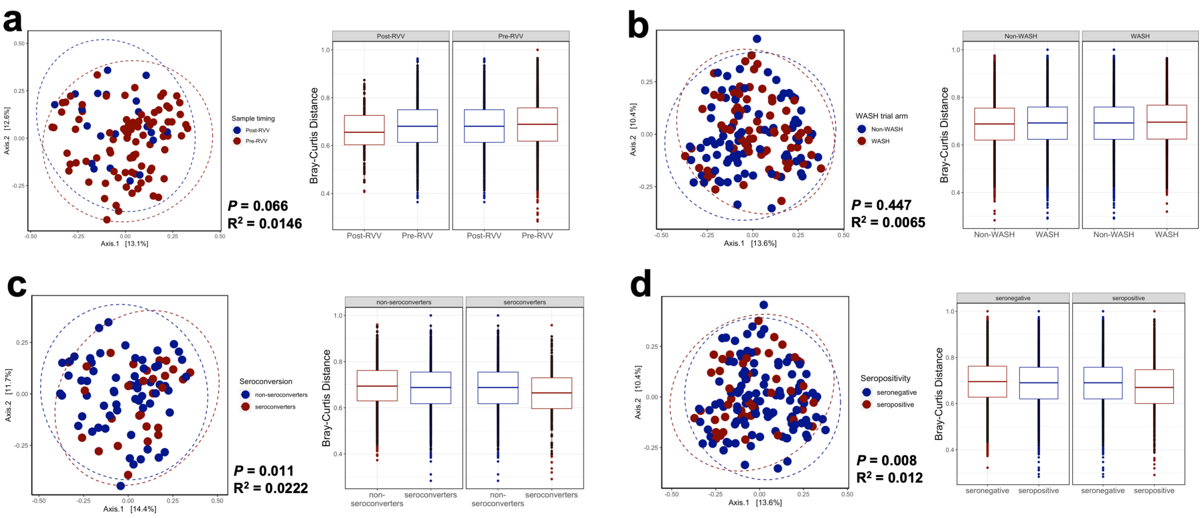


**Fig. S1.** PCoA and associated Bray-Curtis distances to assess beta diversity by seroconversion status between samples collected immediately before or after the first RVV dose (a), by randomized WASH trial arm (b), by seroconversion status in a restricted dataset including samples collected within 14 days of either vaccine dose (c) and by seropositivity status (d) using PERMANOVA analysis.

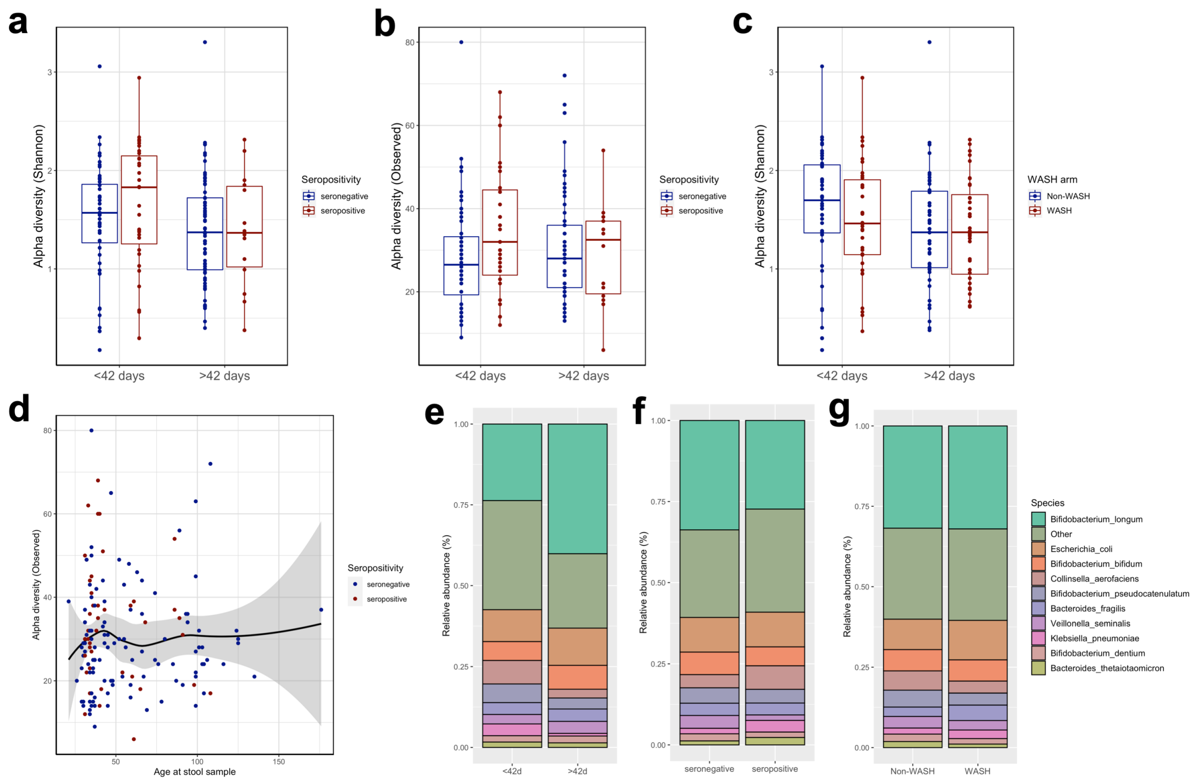


**Fig. S2.** Alpha diversity, as assessed using the Shannon index and number of observed species, between seropositive and seronegative infants (a-b) and by randomized WASH arm (c). (d) Associations between number of observed species and age at stool sample collection. Species composition in seropositive vs seronegative infants (e), early vs late samples (f) and WASH vs non-WASH infants (g).

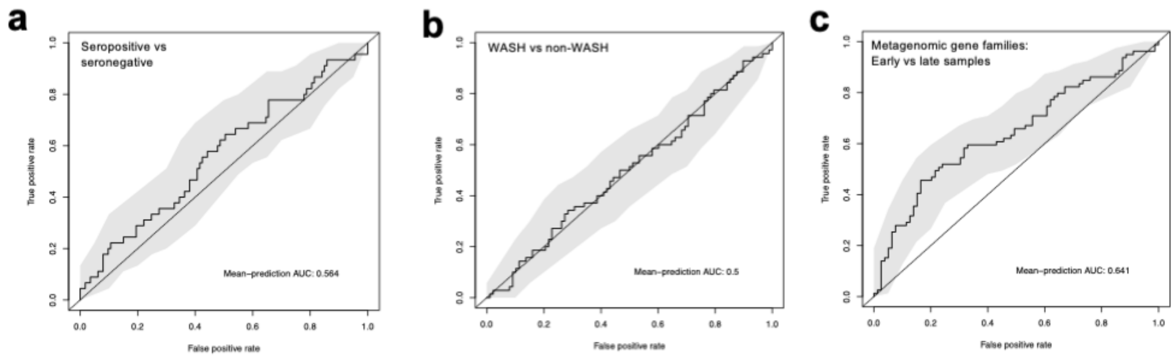


**Fig. S3.** Random forest classification ROC of models using taxonomic data to compare seropositivity status (a) and randomized WASH arm (b), and using metagenomic pathways to compare randomized WASH arm (c), seroconversion status (d), seropositivity status (e) and early vs late samples (f).
